# Mathematical-structure based Morphological Classification of Skin Eruptions and Linking to the Pathophysiological State of Chronic Spontaneous Urticaria

**DOI:** 10.1101/2022.11.04.22281917

**Authors:** Sungrim Seirin-Lee, Daiki Matsubara, Yuhki Yanase, Takuma Kunieda, Shunsuke Takahagi, Michihiro Hide

## Abstract

Chronic spontaneous urticaria (CSU) is one of the most intractable human-specific skin diseases. However, as no experimental animal model exists, the mechanism underlying disease pathogenesis *in vivo* remains unclear, making the establishment of a curative treatment challenging. Here, using a novel approach combining mathematical modeling, *in vitro* experiments and clinical data analysis, we show that the pathological state of CSU patients can be inferred by geometric features of the skin eruptions. Based on our hierarchical mathematical modelling and the analysis of 105 CSU patient eruption pattern geometries, analyzed by six dermatologists, we demonstrate that the eruption patterns can be classified into five categories, each with distinct histamine, basophils, mast cells and coagulation factors network signatures. Furthermore, our network analysis revealed that tissue factor degradation/activation likely determines boundary/area pattern, and that the state of spontaneous histamine release from mast cells may contribute to divergence of the boundary pattern. Thus, our study not only demonstrates that pathological states of diseases can be defined by geometric features but will also facilitate more accurate decision-making to manage CSU in the clinical setting.

## INTRODUCTION

The skin is the largest organ in the human body and plays an important role in maintaining homeostasis and protecting the body from the external environment. Antigens, chemicals, and ultraviolet rays, and endogenous substances can cause cutaneous inflammatory reactions and tumorigenesis. Although certain skin diseases are not life-threatening, such as atopic dermatitis, psoriasis vulgaris, and chronic urticaria, they have a considerable impact on society due to how they affect an individual’s quality of life. Urticaria is one of the most common human skin disorders, affecting at least one in five people during their lifetime (Lee et al. 2017; Maxim et al. 2018). It is characterized by appearance of eruptions on the skin, also known as wheals, that are of various shapes ranging from a few millimeters to several centimeters in size and is typically accompanied by itching, with patients potentially being affected for years to even decades. Among the different subtypes of urticaria, chronic spontaneous urticaria (CSU) is characterized by the spontaneous appearance and disappearance of local skin edema and flares (Zuberbier et al. 2022). The pathophysiological states associated with CSU includes autoimmune responses, cellular infiltrates, coagulation, and the complement system (Kolkhir et al. 2022). However, how these responses contribute to wheals remains unknown, which is largely due to the limited understanding and scarcity of clinical data on the *in vivo* pathological structure of urticaria. Furthermore, due to the human-specific nature of the disease, there is also lack of an appropriate animal model system that can recapitulate the entire disease pathology. Consequently, elucidating the underlying pathophysiological dynamics *in vivo* remains challenging, and developing patient-specific effective treatments for urticaria remain difficult.

To overcome the limitations of the lack of human-specific skin disease models, we developed a novel approach linking the geometric features of skin eruption with pathophysiological structures *in vivo*. As all skin diseases are characterized by “visible outputs”, skin eruptions can be considered as a reflection of *in vivo* pathophysiological dynamics. However, eruption geometry has neither been fully understood nor explored as a measure of the differences in pathological states between patients. Thus, in this study, we focused on the morphological features of skin eruptions and developed a novel approach linking the eruption geometry of patients to the *in vivo* pathological dynamics of CSU using hierarchical mathematical modeling, *in vitro* experiments, and clinical data analysis of the eruption patterns of urticaria. We show that the major *in vivo* pathophysiological state of CSU can be typically defined as per one of five skin eruption pattern types, thereby also demonstrating the potential clinical applicability of this strategy in developing as a new diagnostic and treatment methods in a clinical setting.

## RESULTS

### CSU mathematical modelling based on pathophysiological structures

To design our CSU mathematical model, we first defined all the necessary pathophysiological molecules and their interactions that have been associated with CSU, based on *in vitro* experimental studies (Fig. 1A). Firstly, we took into consideration that external stimuli or internal auto-allergens and auto-antibodies stimulate both basophils and mast cells to release mediators by crosslinking the high-affinity IgE receptors (FcχRI) on their surfaces (Karasuyama et al., 2009, Stone et al., 2010, Kaplan et al., 2022) [Fig. 1A (0)]. Next, that this process subsequently enhances TF expression on vascular endothelial cells [Fig. 1A (1)], which induces the extrinsic coagulation cascade (Fomby et al., 2010, Mackman, 2009, Hide & Kaplan, 2022) [Fig. 1A (2_a_)]. Consequently, the coagulation factors increase degranulation of basophils [Fig. 1A (2_b_)] or skin mast cells via gap formation by vascular endothelial cells [Fig. 1A (2_a_), (3), (4)] leading to edema [Fig. 1A (5), (6)] (Yanase et al., 2021). Furthermore, adenosine, which is simultaneously released following degranulation of both human peripheral basophils and skin mast cells (Rudich et al., 2012), suppresses histamine release and TF expression (Matsuo et al., 2018, Morioke, et al., 2018), we thus also took this into consideration (Fig. 1A, green inhibition symbols). Based on these facts, we applied the pathophysiological structure into a network structure (Fig. 1B), focusing on the following main factors that are expected to have a key role in the *in vivo* network: 1) concentration of histamine released from basophils ([*H*_*B*_]), 2) TF expression on vascular endothelial cells ([*TF*]), 3) coagulation factors leaked from blood vessels ([*C*]), and 4) histamine released from mast cells ([*H*_*M*_]).

**Fig. 1.**
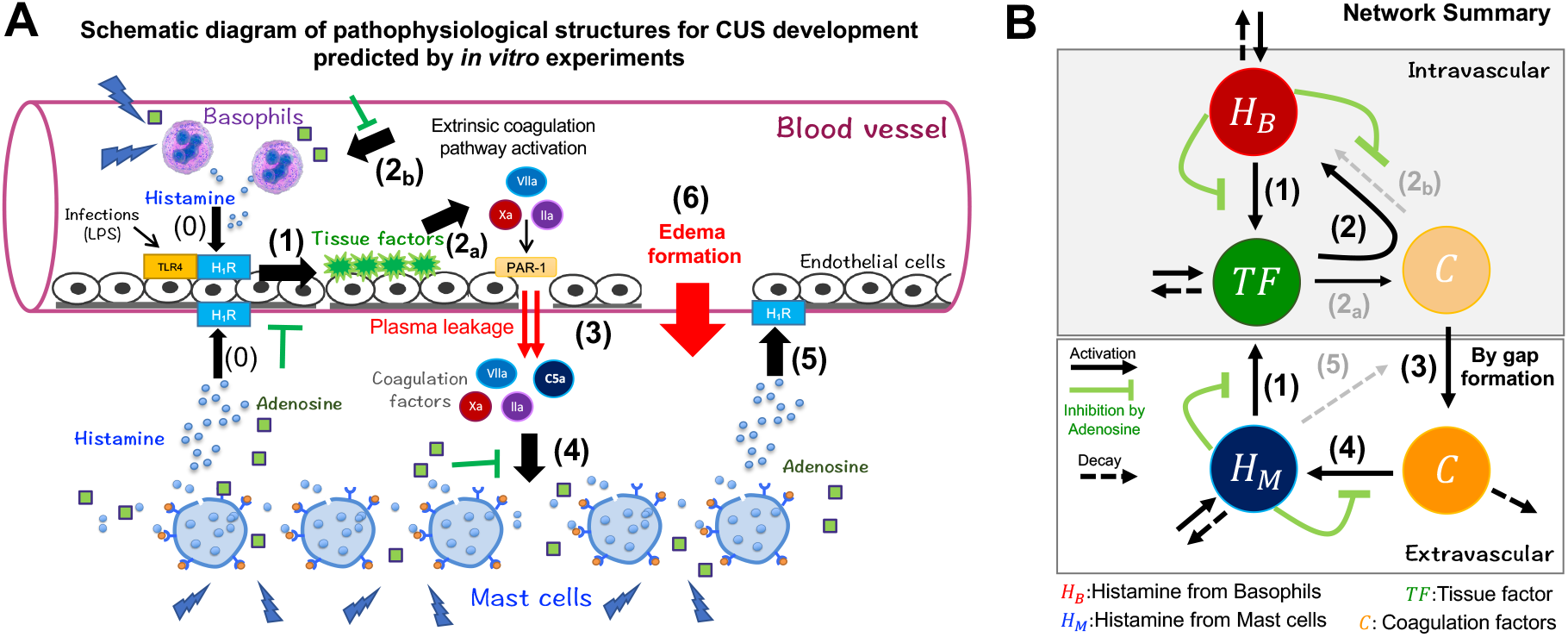
*in vivo* pathophysiological network of chronic spontaneous urticaria (CSU). (A) Schematic diagram of CSU development predicted using *in vitro* experiments. Number labels correspond with those in (B). Black arrows and green symbols indicate positive (activation) and negative (suppression/inhibition) regulation, respectively. The order of pathophysiological progression is sequentially numbered. (B) Summary of the network based on the description in (A). Networks that do not appear explicitly in the mathematical model are depicted by gray dotted arrows. In the mathematical model, arrows (2_a_) and (2_b_) were simplified to solid arrows (2). Networks (2_a_) and (5) were integrated into the modeling of the gap-forming network (3).

We developed the hierarchical mathematical model integrating both the intravascular and extravascular dynamics using *in vitro* experimental data. The dermis of the skin can be considered as a thin two-dimensional (2D) sheet with numerous capillaries spread over it. We constructed a 2D spatial model with the intravascular space distributed homogeneously throughout the dermis. Based on the network (Fig. 1B), we constructed the following model (see Supplementary Notes 1.1−1.5 for details on model construction and parameters estimation).

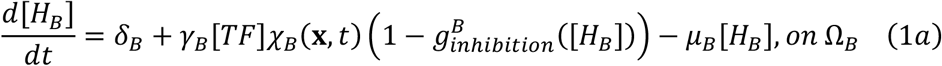

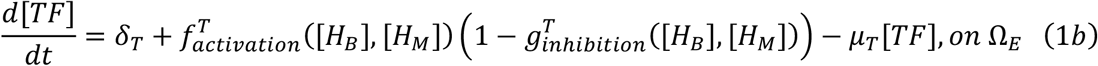

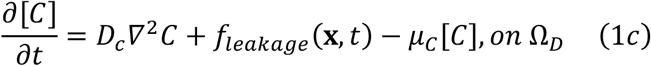

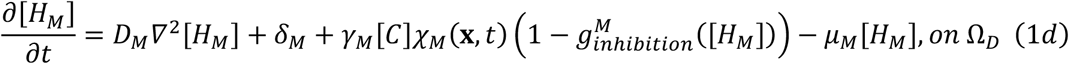

where, *Ω*_*D*_(⊂ ℝ^2.^) is the dermal region, *Ω*_*E*_ is the vascular endothelial tissue, and *Ω*_*B*_(⊂ *Ω*_*E*_) is a randomly chosen local region of vascular endothelial tissue where the basophils adhere to the endothelial cells. Furthermore, *δ*_*B*_, *δ*_*T*_, and *δ*_*M*_ are the basal production rates and *µ*_*B*_, *µ*_*T*)_, and *µ*_*M*_ are the basal decay rates. *γ*_*B*_ and *γ*_*M*_ are histamine release rates of basophils and mast cells, respectively (network (2) and (4) in Fig. 1A, B). The terms *χ*_*B*_ (***x***, *t*) and *χ*_*M*_(***x***, *t*) are step functions, assumed to be 1 when each basophil and mast cell contained histamine and 0 when they each released their total histamine content (Hattori & Seifert R, 2017; Seirin-Lee et al., 2020). We quantitatively estimated the main network functions, 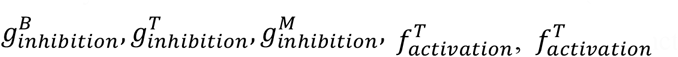, and *f*_*leakage*_(***x***, *t*) based on the *in vitro* experiment data.

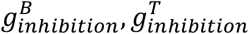 and 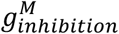 represent the inhibition rates of adenosine which is produced by adenosine triphosphate (ATP) released simultaneously with and in proportion to histamine from basophils and mast cells (Rudich et al., 2012) (green inhibition symbols in the networks shown in Fig. 1A, B). Adenosine is known to suppresses the histamine release of human peripheral basophils and human skin mast cells (hsMCs) in response to anti-IgE in a concentration-dependent manner (Matsuo et al., 2018). Based on our previous data (Matsuo et al., 2018), we directly analyzed the inhibitory rates of histamine released from basophils and hsMCs via adenosine (Fig. 2A and B, respectively). Using these data, we estimated the specific forms of 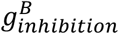 and 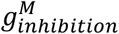 as shown in Fig. 2C:

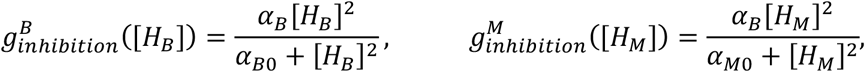

where *α*_*B*_ and *α*_*B*_ are the maximum inhibition rates, *α*_*B*0_ and *α*_*M*0_ are the constants that determine the degree of increase in the curves.

**Fig. 2.**
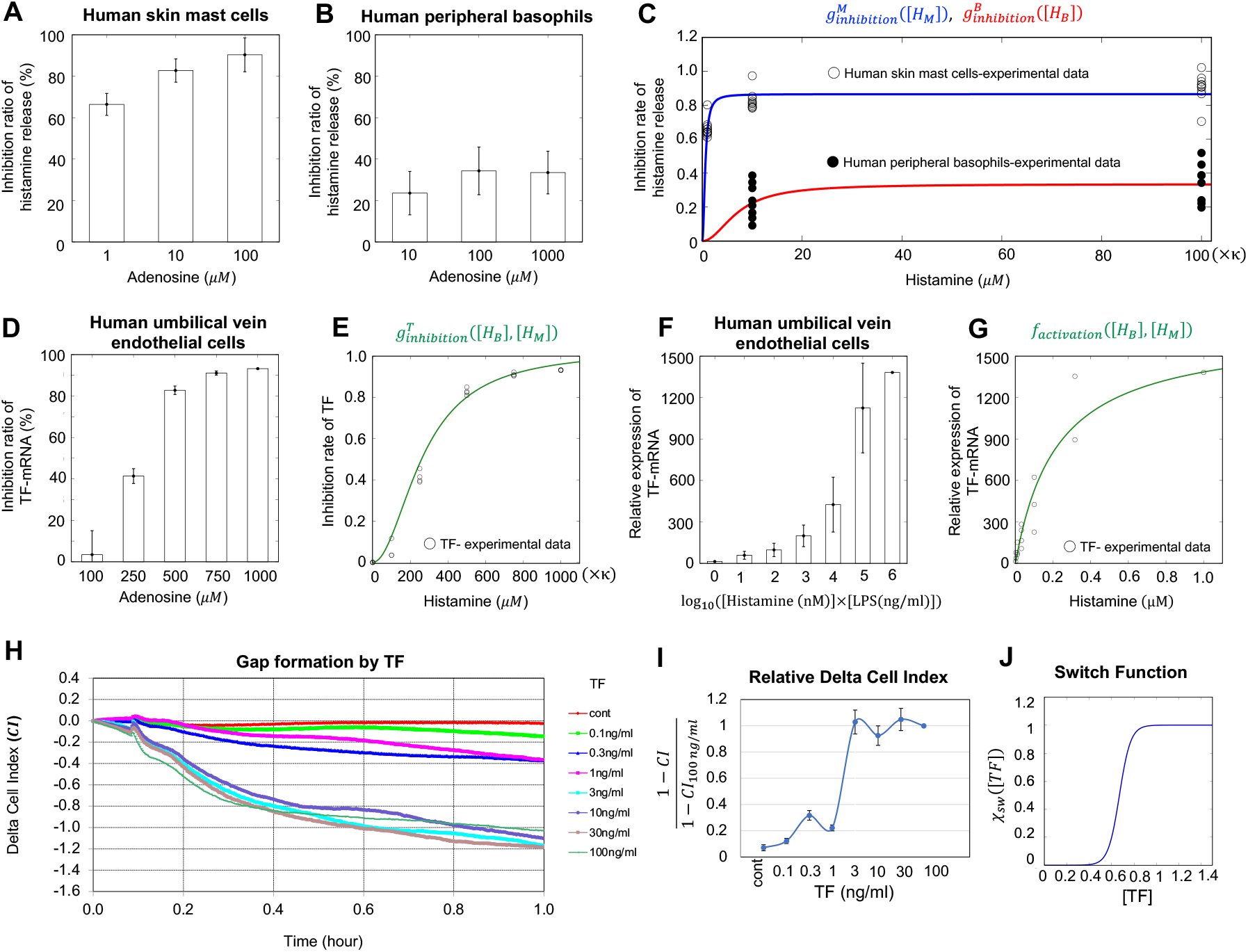
*in vitro* experiments for key networks and estimation of model functions. (A, B) Inhibition ratio of histamine release from human skin mast cells (hsMCs)/human peripheral basophils in response to adenosine, calculated from data of Matsuo et al., 2018. (C) Inhibitory functions estimated from data of (A, B). (D) Inhibition ratio of tissue factor (TF) mRNA from HUVECs induced by adenosine, calculated from experimental data of Yanase, Morioke, et al., 2018. (E) Inhibitory function estimated from the data of (D). (F) Relative expression of TF mRNA from HUVECs induced by co-stimulation with histamine and LPS. Data used were from Yanase, Morioke, et al., 2018. (G) Activation by TF estimated from data of (F). κ is given in equation (3) and 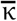 is a scaling parameter that converts LPS concentration to histamine concentration (details are presented in Supplementary Notes 1.2-1.3). (H) TF concentration-dependent gap formation. Temporal variation in cell index (CI) at different TF concentrations. Endothelial cells were stimulated with TF at indicated concentrations in Dulbecco’s modified Eagle’s Medium (DMEM)/F12 with 2% bovine serum albumin (BSA) and 1% factor VIII-deficient human plasma without any supplements and fetal calf serum (FCS). (I) Relative scale of CI, which was scaled by data obtained at 100 ng/mL each time. (J) Estimated switch function, *χ*_*sw*_([*TF*]), obtained by quantitative analysis of variation in CI at different TF concentrations.

Next, using *in vitro* experiment data of the tissue factor (TF) suppression by adenosine (Yanase, Morioke et al., 2018), we also analyzed the inhibitory rates of TF mRNA expression based on adenosine concentrations (Fig. 2D) and estimated the inhibitory rate function of TF follows (Fig. 2E):

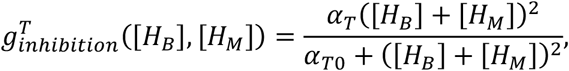

where we estimated the parameter values, the maximum inhibition rate (*α*_*T*_) and a constant (*α*_*T*0_), by the least squares method.

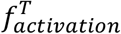, represents a function of histamine-induced TF expression (networks (1) in Fig. 1A, B). TF in human umbilical vein endothelial cells can be induced by histamine and synergistically increased by co-stimulation with histamine and lipopolysaccharide (LPS) (Yanase, Morioke et al., 2018). To reflect the potential synergistical output of TF activation in our model, we directly analyzed previous experimental data on the relative expression level of TF mRNA induced by co-stimulation with histamine and LPS (Fig. 2F). We then converted the result to data based on the histamine-only concentration and estimated the TF activation function as follows (Fig. 2G):

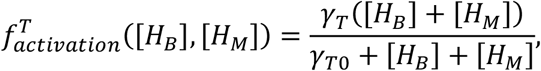

where *γ*_*T*)_ is the maximum activation rate and *γ*_*T*0_ is a constant that determines the degree of increase in the curve.

The TF expressed on endothelial cells induces the extrinsic coagulation cascade (Fomby et al., 2010; Mackman, 2009), and activated coagulation factors produced by the extrinsic coagulation pathway induce intracellular gap formation between endothelial cells via PAR-1 (Yanase, Takahagi, et al., 2018). Thus, the coagulation factors leaked from intravascular, namely, *f*_*leakage*_ (***x***, *t*) can be determined by the dynamics of intercellular gap formation.

In order to determine the detailed form of *f*_*leakage*_ (***x***, *t*), we performed *in vitro* experiments using human umbilical vein endothelial cells (HUVECs). We looked for the changes in the adhesion area and morphology of the cells cultured on the electrodes with varying TF and histamine concentrations, respectively. We first measured the time-related cell index (CI) with varying TF by using an impedance analysis (Fig. 2H). We found that the CI was not significantly changed following treatment with TF at < 1 ng/mL, whereas it was dramatically decreased with a TF concentration > 3 ng/mL. Notably, the change in CI was classified at two levels and its dynamics was switched by a threshold concentration of TF between 1–3 ng/mL. This result suggests that TF plays a role in inducing the leakage of coagulation factors via switch-like gap formation.

Based on this experimental result, we inferred the TF-dependent switch-like function, *χ*_*sw*_([*TF*]), by calculating the relative CI, (1 − *CI*)/(1 – *CI*_100*ng*/*ml*_), which gives the relative variation in the size of the CI for each time point. We calculated the average relative CI at [0.2, 0.6] (hour) for each TF concentration and found the switch-like function as shown in Fig. 2I and 2J. Furthermore, we found that the rate of gap area expansion is determined in a concentration-independent manner for both TF and histamine (Supplementary Note 1.5, Fig. S1 in mathematical and experimental details). Taken together, we obtained

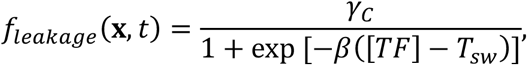

where *γ*_*C*_ is a leaked rate of coagulation factors (Fig. 2J).

At this stage, we had completed the CSU mathematical model (1a)–(1d) based on experimental analysis of the key pathophysiological networks. The eruptions of CSU visibly appeared on the skin where the leaked coagulation factors from blood vessel or histamine concentration were high in the dermis. Therefore, we plotted the eruption state of the skin using the eruption state function (defined in the Online Methods).

### Recapitulation of eruption patterns in urticaria using a mathematical model

We next validated whether our mathematical model could successfully recapitulate the CSU eruption patterns. Depending on the parameter values, we identified several eruption patterns observed in the patients with urticaria (Fig. 3A–E, Fig. S2). Based on geometric characteristics, we categorized the different eruption patterns identified in our mathematical model as follows: annular, broken-annular, geographic, circular, and dot patterns.

**Fig. 3.**
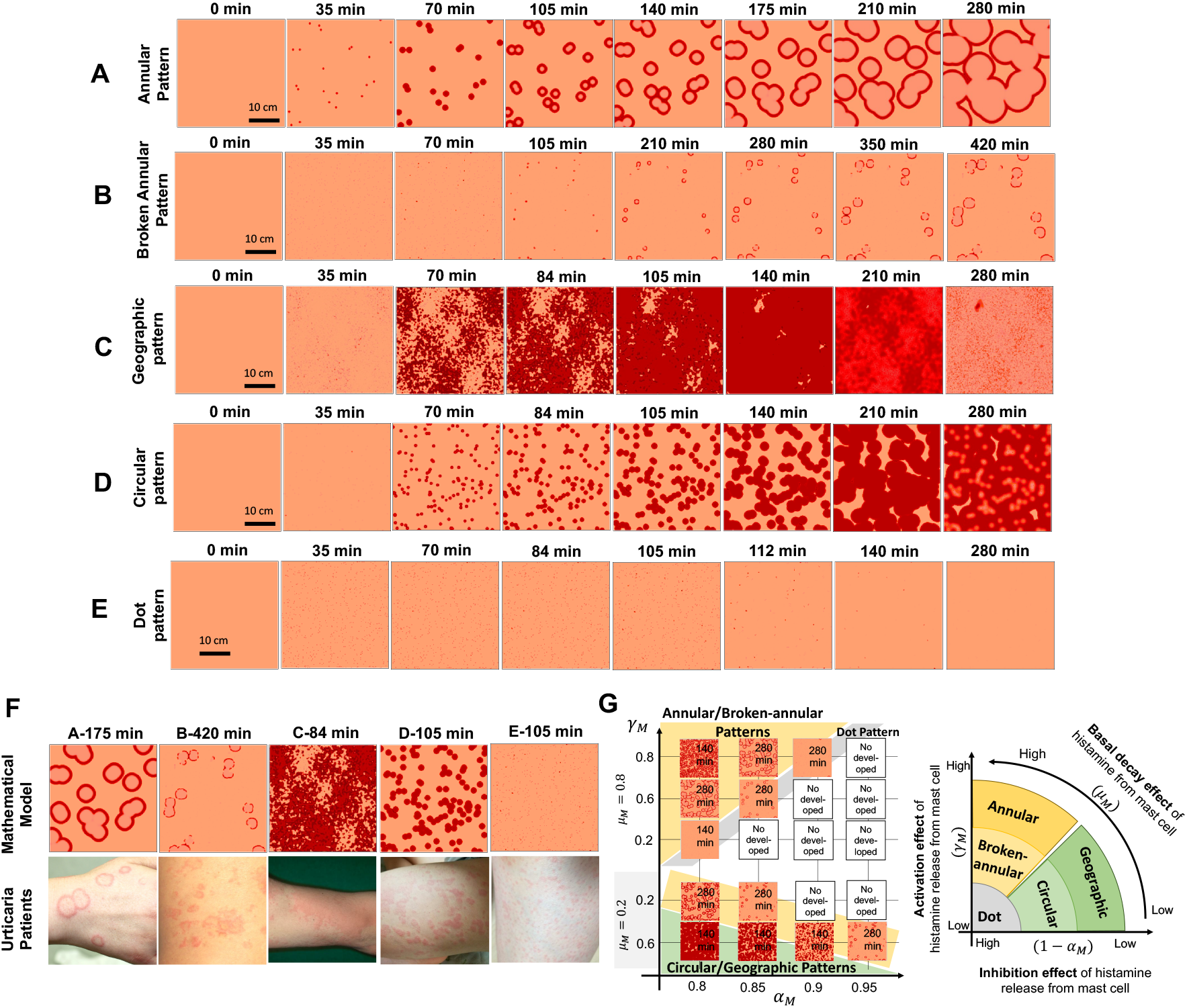
Five wheal patterns identified using a mathematical model. (A–E) Evolution of each wheal pattern. (F) Comparison of wheal pattern using a mathematical model at a time point with that of urticaria patients. Details of parameter values are shown in Tables S1. (G) Left panel shows eruption patterns by changing parameter spaces: activation of histamine release (*γ*_*M*_), decay rate of histamine (*µ*_*M*_), and inhibition rate of histamine (*α*_*M*_) from mast cells. Right panel shows a schematic diagram of eruption pattern based on three parameters.

The eruption image data of patients in clinical medicine represent the data of a certain time point of eruption development dynamics. Thus, we compared the *in silico* eruption patterns at some fixed time point with the image data collected for CSU patients who visited the hospital (Fig. 3F). We found that patterns among the patient data were highly similar to all pattern types observed in the *in silico* experiments, hence validating our CSU mathematical model.

### Classification of eruption geometry in urticaria patients according to eruption geometry criteria

Interestingly, during the *in silico* validation of our mathematical model, we observed that eruption patterns were typically identified as one of five types when parameter values were varied (Fig. 3G). Therefore, we next sought to determine whether all eruption patterns appearing in CSU patients could be classified into specific types and then linked to *in vivo* pathologies. To this end, we first developed classification criteria defining the clinical features of the eruption (wheal) geometry found in the *in silico* patterns (Table 1). We designated the resultant table of clinical features *Criteria for Classification of Eruption Geometry* (EGe criteria). We classified the patient eruption patterns into two large classes, boundary and area patterns, which we then further classified into five specific patterns, namely annular, broken-annular, geographic, circular, and dot patterns, as observed in the *in silico* experiments.

**Table 1:**
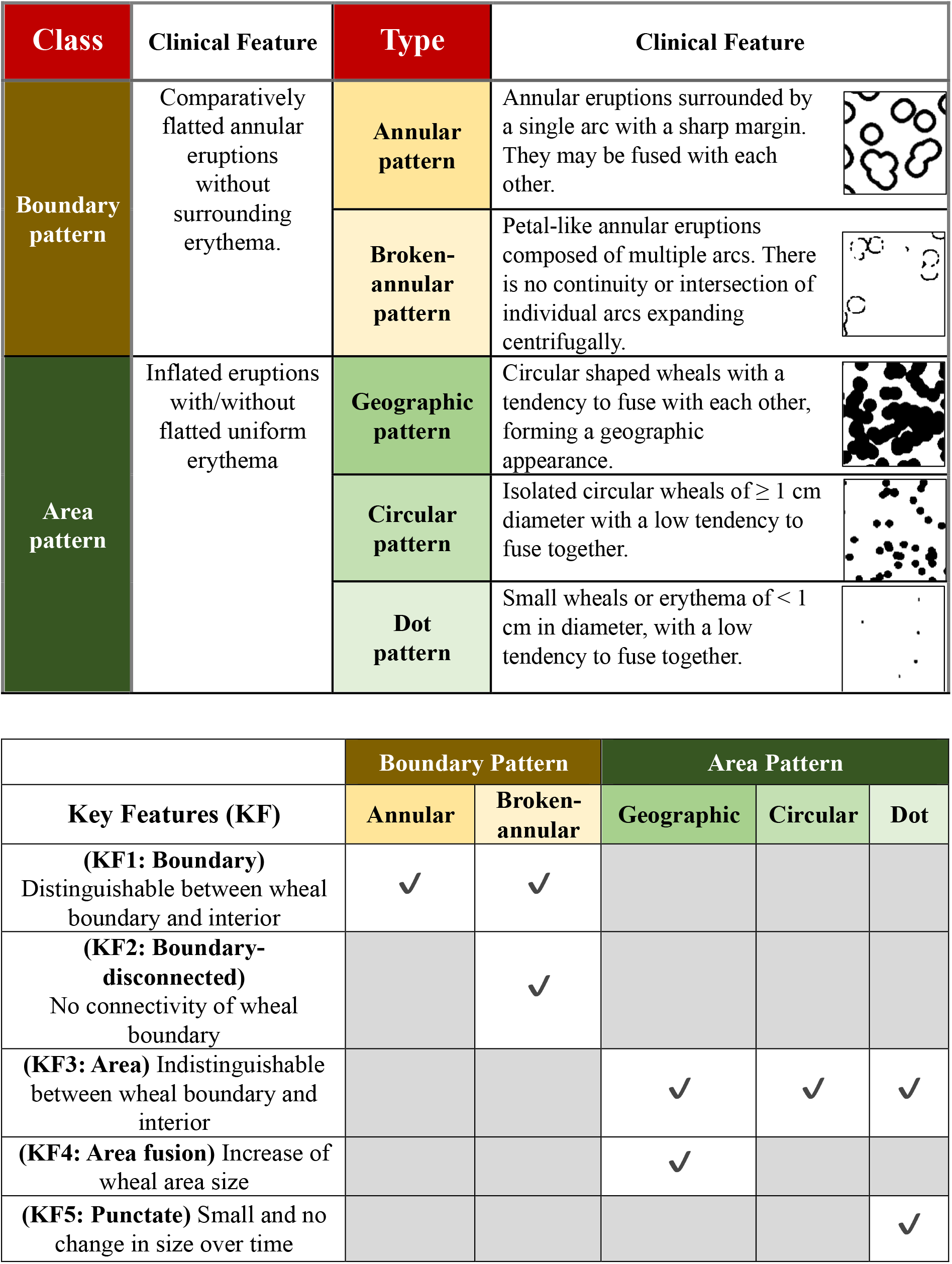
Criteria for classification of eruption geometry (EGe criteria) and summary table of eruption geometries by key features. Morphologically, eruptions in chronic spontaneous urticaria (CSU) are divided into two classes and five subtypes. The below table provides simple means of classifying eruption geometry (EGe) based on five key features.

| Class | Clinical Feature | Type | Clinical Feature |
| --- | --- | --- | --- |
| Boundary pattern | Comparatively flattened annular eruptions without surrounding erythema. | Annular pattern | Annular eruptions surrounded by a single arc with a sharp margin. They may be fused with each other. |
|  |  | Broken-annular pattern | Petal-like annular eruptions composed of multiple arcs. There is no continuity or intersection of individual arcs expanding centrifugally. |
| Area pattern | Inflated eruptions with/without flattened uniform erythema | Geographic pattern | Circular shaped wheals with a tendency to fuse with each other, forming a geographic appearance. |
| | | Circular pattern | Isolated circular wheals of $\geq 1$ cm diameter with a low tendency to fuse together. |
| | | Dot pattern | Small wheals or erythema of $< 1$ cm in diameter, with a low tendency to fuse together. |

|  | Boundary Pattern |  | Area Pattern |  |  |
| --- | --- | --- | --- | --- | --- |
| Key Features (KF) | Annular | Broken-annular | Geographic | Circular | Dot |
| **(KF1: Boundary)**<br>Distinguishable between wheal boundary and interior | ✓ | ✓ |  |  |  |
| **(KF2: Boundary-disconnected)**<br>No connectivity of wheal boundary |  | ✓ |  |  |  |
| **(KF3: Area)** Indistinguishable between wheal boundary and interior |  |  | ✓ | ✓ | ✓ |
| **(KF4: Area fusion)** Increase of wheal area size |  |  | ✓ |  |  |
| **(KF5: Punctate)** Small and no change in size over time |  |  |  |  | ✓ |

To confirm whether the eruption patterns of actual CSU patients could be classified into one of the eruption types in the EGe criteria, we collected the eruption image data of 105 patients who visited the Hiroshima University Hospital with CSU-related skin eruptions (see Online Method). The data were then analyzed by six dermatologists, based on the EGe criteria. We evaluated the reliability **SB (**≡[number of doctors who selected the sample as one of the types in the EGe criteria]/[number of dermatologists (6)]), of the statistical result using the vote-counting method.

The results showed that 87.6% of the samples could be classified as one of the eruption types with 100% **SB** reliability (Fig. 4A, left panel), and 0% of the samples was unclassifiable, with a reliability **SB** > 83% (Fig. 4A, right panel). We also statistically confirmed that the classifiable rate of 87.6% is not a random chance by the *P*-value (Supplementary Note 1.6). This result proves that the eruption types in CSU patients could be classified into one of the patterns of the EGe criteria developed on the basis of our mathematical model. This suggests that the eruption dynamics of CSU patients can be determined using our CSU mathematical model.

**Fig. 4.**
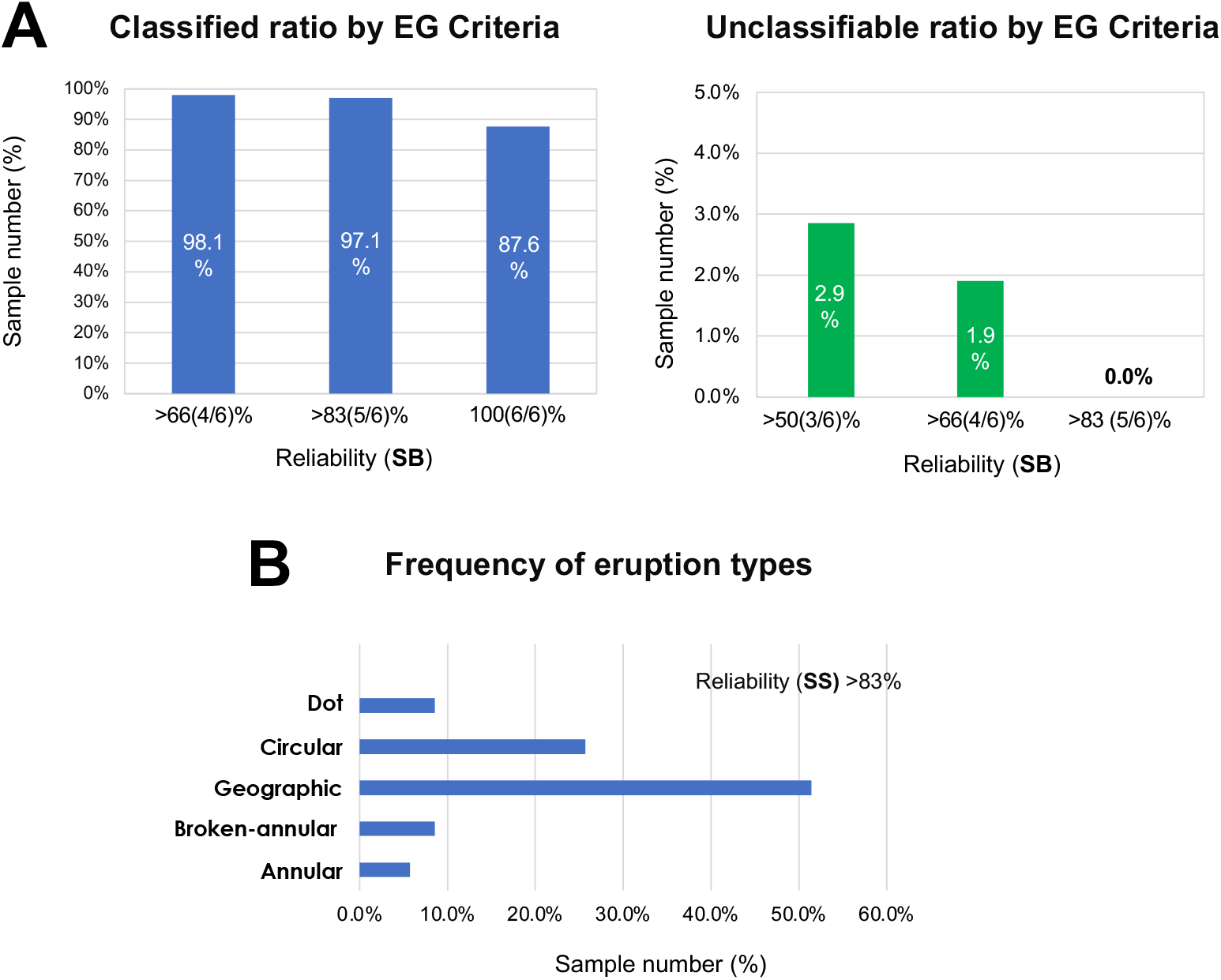
Classification of eruption patterns by EGe criteria for CSU patients. (A) Ratio of patient samples in which six dermatologists selected one of the pattern types defined by the EGe criteria (left panel); ratio of patient samples selected as not classifiable by the EGe criteria (right panel). Data was calculated based on reliability **SB**. (B) Frequency of patient eruption types. Data were selected with reliability **SS** > 83%.

Next, we investigated which types of eruption patterns were commonly observed in CSU patients. To obtain results with a higher reliability, we introduced a new measurement of reliability (**SS**) defined as: **SS** = [**SB** of type selection]×[**SB** of class selection] (Fig. 4A, Online Methods). From the analyzed data with the reliability **SS**, we found that geographic and circular types were observed at high frequencies, whereas boundary patterns appeared fewer times than area patterns did.

### Linking the key features of eruption geometry to related pathological networks via mathematical model

We next extracted the minimal sets of key characteristics of eruption patterns from the features defined in the EGe criteria (Table 1): distinguishable (KF1)/indistinguishable (KF3) between the boundary and interior of the wheal, disconnectedness in the wheal boundary (KF2), fusion of the wheals (KF4), and no change in size of wheals (KF5). This showed show that each of the eruption patterns could be identified using appropriate combinations of the five features. Specifically, the boundary pattern is identified by KF1, the annular/broken-annular pattern is identified by KF1 and KF2, the geographic/circular pattern is identified by KF3 and KF4, and the dot pattern is identified by KF3 and KF5.

We then looked to see how each eruption pattern is connected to the network of pathological dynamics. Thus, we performed a global sensitivity analysis of the Extended Fourier Amplitude Sensitivity Test (eFAST) (Saltelli et al., 1999; explained in detail in Supplementary Note 1.7), which tells us which networks play an important role for a given sensitivity function. We defined the sensitivity function based on the five key features (shown in Table 1). Specifically, we defined the five sensitivity functions to quantitatively measure the degree of alteration of the features of each eruption pattern in response to changes in parameter values representing the network intensity (Fig. 5, Supplementary Note 1.7 shows the details of sensitivity functions). Thus, this analysis identifies the networks that play an important role in each eruption pattern.

**Fig. 5.**
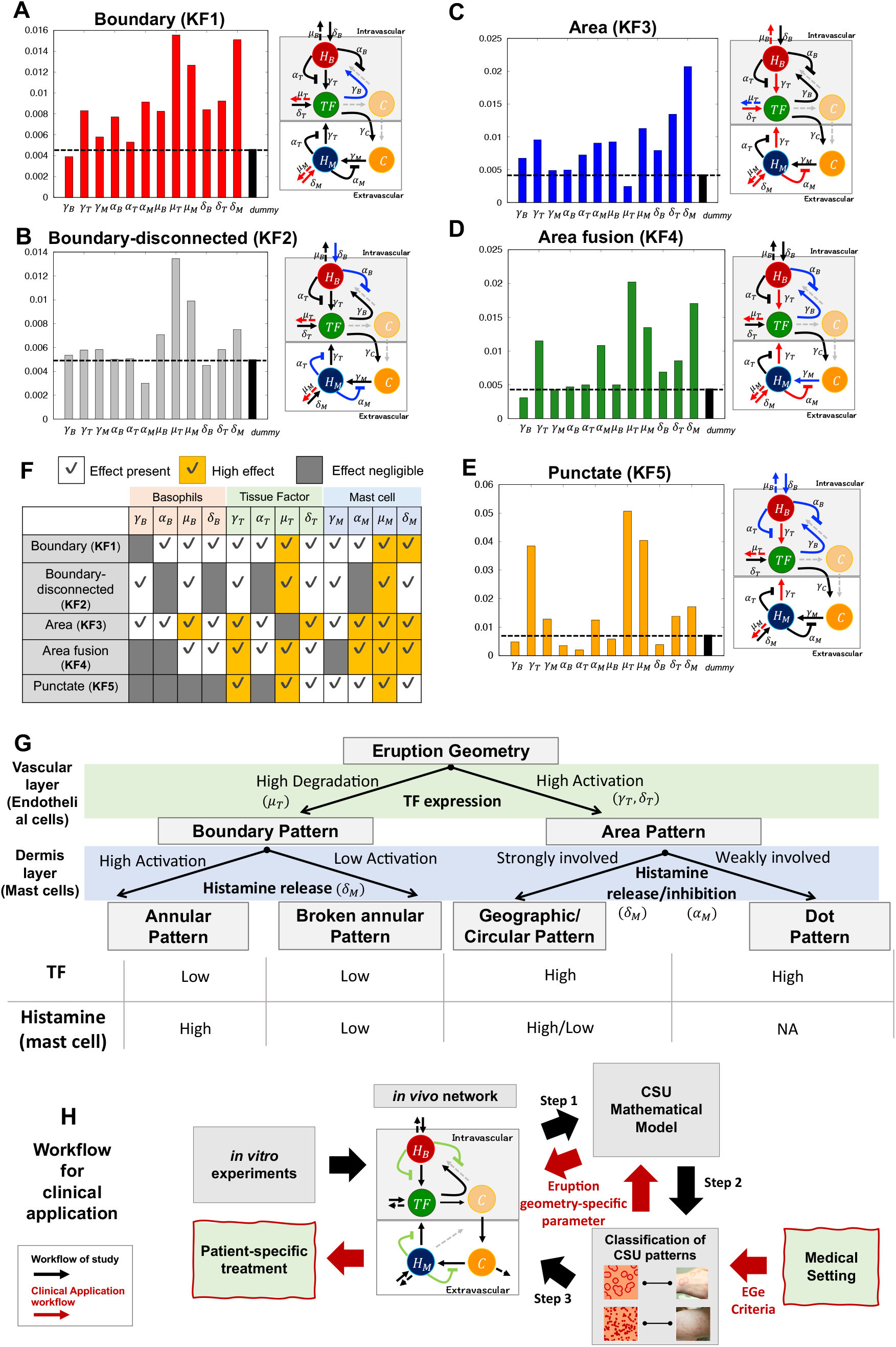
Linking key features of wheal geometry to related networks. (A–E) Results of the first index determined using Extended Fourier Amplitude Sensitivity Test (eFAST) sensitive analysis of each feature. The first order index of the dummy parameter was calculated to confirm that index values in panels A–E yielded reliable data to determine the significance of model parameters (Marino et al., 2008). For model parameters with an index less than or equal to that of the dummy parameter, the parameters are considered not significantly different from zero. Networks with indexes lower than (negligible effect) and double the height of the dummy parameter (high effect) are indicated in blue and red, respectively. (F) Summary table of sensitivity analysis. (G) Summary of analysis results of the pattern types and main network. The table at the bottom shows relative levels of tissue factor (TF) expression and histamine released by mast cells for each eruption pattern. (H) Summary of workflow and clinical application. Black arrows show the study workflow and red arrows represent the general clinical applications of our research, leading to potential patient-specific treatments for intractable skin diseases.

We first noted that most key features had a relatively high association with the networks related to TF and mast cells (Fig. 5F, yellow-colored regions), suggesting that the dynamics of TF and mast cell may be important players in determining the geometrical property of CSU eruptions. We next compared the difference between boundary and area features. In the boundary feature (KF1), the decay rate of TF (*µ*_*T*_) highly influenced the features (Fig. 5A, F). By contrast, we found that *µ*_*T*_ had a negligible effect on area feature (KF3) (Fig. 5C, F). In addition, the activation network of TF (*γ*_*T*_ and *δ*_*T*_) had a considerable effect on the area feature but not on the boundary feature, although an effect was evident (Fig. 5F). These results suggest that the feature of boundary/area pattern is likely to be highly involved in TF degradation/activation.

Next, to determine the network that played a critical role in the divergence of the annular and broken annual patterns, we compared the sensitive analysis result between the boundary (KF1) and boundary disconnectedness (KF2) features (Fig. 5 A, B). We found that the effect of the basal activation network (*δ*_*M*_) of histamine release from mast cells in the disconnectedness feature (KF2) was not as high as that of the boundary feature (KF1), indicating that the state of spontaneous histamine release from mast cells may be the critical factor to make the divergence of the annular and broken annual patterns.

We then compared the area fusion (KF4) and punctate (KF5) features (Fig. 5D, E). The first notable differences between these two features were the activating effect of spontaneous histamine release from mast cells (*δ*_*M*_) and the inhibitory effect of histamine release from mast cells (*α*_*M*_). Both the networks (*δ*_*M*_ and *α*_*M*_) showed a high effect on the area fusion feature, whereas they exhibited relatively negligible effects on the punctate feature. This result suggests that the area types of eruption patterns may become geographic/circular type or dot type depending on whether the histamine dynamics from mast cells are strongly involved or not. We summarized the classification of eruption types and their relations with TF and histamine dynamics of mast cell based on the sensitive analysis in Fig. 5G.

## DISCUSSION

In this study, we established a novel approach to overcome the limitations of clinical data by developing a three-step research strategy: generating a mathematical model that recapitulates eruption patterns from the pathological structures (Fig. 5H, Step 1), establishing EG criteria for the clinical classification of eruptions in patients based on *in silico* eruption patterns (Fig. 5H, Step 2), and linking the features of eruption geometry to critical networks of pathological structures (Fig. 5H, Step 3). With this this stepwise approach, we determined the *in vivo* pathological molecular dynamics in urticaria patients via their eruption geometry, by extracting the geometric features of skin eruptions from patients with urticaria and linking these features to the parameter values of the CSU mathematical model.

In the first step, we successfully developed a hierarchical urticaria mathematical model in which all the model equations were verified by *in vitro* experiments. To our knowledge, this is the first study wherein the pathophysiological steps involved in the development of CSU was structurally proven *in silico*. The first conceptual mathematical model of urticaria was proposed in our recent work (Seirin-Lee et al., 2020). In the previous model, we proposed that the inhibitory regulation of histamine release from mast cells plays an important role in creating the multifarious patterns of the wheal geometry, although this finding has not been experimentally verified. In this study, the inhibition dynamics of adenosine was modeled based on the experimental data (Matsuo et al., 2018) and proved that the inhibitory regulation in mast cells is likely to be involved in the classification of eruption types with interactions in other physiological networks (Fig. 4G, Fig. 6). Furthermore, our CSU model proposes that the hierarchical structure of urticaria development as well as the key players, such as TF and histamine released from mast cells, are critical in generating diverse eruption patterns and determining the eruption types.

In the process of developing the mathematical model, we also found that the intracellular gap in endothelial cells of the blood vessels is formed at a threshold level that depends on TF concentration. This finding suggests that urticaria may develop only when stimuli are presented at a level above a certain threshold. Thus, the gap formation dynamics based on TF switch may affect not only the formation of eruption patterns but also the onset of urticaria.

Using our model, we also identified five eruption patterns and created EGe criteria for the classification of urticaria eruptions in real patients. Application of the EGe criteria confirmed that patients with CSU could be classified into one of the five types, and the types included in the area pattern were observed more frequently than those in the boundary pattern in the patients. While boundary patterns are observed specifically in patients with spontaneous urticaria, area patterns are basic wheal structures seen in both spontaneous and inducible types of urticaria. Therefore, area patterning may reflect the ubiquitous molecular mechanisms common to patients and various types of urticaria, thereby resulting in a high patterning frequency. Taken together, these results indicate that our EGe criteria could be a tool of practical approach for clinicians to apply in determining eruption patterns of patients in future.

In the final step of our research strategy, we inferred the pathophysiological networks that are critically involved in each feature of the eruption geometry. We found that the balance of effects from TF and the histamine from mast cells play a critical role in determining the types of eruption patterns. Our result suggests that the level of TF expression may be involved in an underlying mechanism mediating the creation of notable differences in eruption shapes such as boundary or area patterns. Furthermore, the kinetics of histamine released from mast cells may also play a role in determining the specific type of the eruption that develops in different patients. This implies that even suppressing the action of a specific network using a drug may not always exhibit the same effectiveness among patients exhibiting eruptions with different shapes. In fact, antihistamines are effective for 70–80% of CSU patients (Maurer et al. 2010), and certain patients refractory to antihistamines, respond well to anticoagulants and protease inhibitors (Chua et al. 2005, Parslew et al. 2000, Takahagi, Shindo et al. 2010). Thus, our approach, which proposes a pathophysiological *in vivo* network based on the feature of eruption geometry, is expected to propose a patient-specific treatment based on the skin eruption geometry (Fig. 5H, red arrows).

Urticaria involves both temporal and spatial dynamics, and the eruptions show highly dynamic transitions from the phases of eruption emergence, development, and disappearance. Our study focused only on the emergence and development phases and did not consider the disappearance phase. In the current model, the eruptions would continue to expand if the simulation space is not limited. This suggests that the disappearance phase may be caused by a network independent of the onset phase. Understanding the overall dynamics of CSU from the onset to the disappearance phase will be crucial to understand the exact pathological state of patients based on the eruption state. To achieve this, the time-related historical data of patients will need to be acquired and added to our current mathematical model.

In this study, we revealed the association between the area pattern and high TF activation, and between eruption type and kinetics of histamine solely by theoretical analysis. However, the role of TF activation on area patterns rather than boundary ones may be experimentally confirmed by examining levels of coagulation biomarkers such as PF1+2 and D-dimer (Asero et al. 2008, Takahagi, Mihara et al. 2010). Therefore, the relation of wheal pattern to the biomarkers of coagulation as well as autoimmunity will need to be evaluated in the future.

The findings of this study suggest that the skin eruption geometry is linked to the state of *in vivo* molecular networks involved in the pathogenesis of CSU. This study proposes that the understanding of the pathological state of each patient may be enhanced by evaluating the eruption geometry. Furthermore, with an inference of the detailed parameter values for each patient it may be possible to apply our mathematical system to decide patient-specific treatments. Our multidisciplinary approach linking the skin eruption geometry and *in vivo* dynamics could help develop a novel strategy for dermatological research to manage intractable skin disease in an integrated manner.

## Data Availability

All data produced in the present study are available upon reasonable request to the authors

## Abbreviations

(CSU): Chronic spontaneous urticaria
(EGe Criteria): eruption geometry criteria
(TF): tissue factor

## Acknowledgments

We thank Drs. Akiko Kamegashira, Satoshi Morioke, and Akio Tanaka at Hiroshima University Hospital for their performing the classification of the morphology of wheals in the clinical part of the study. We thank Dr. Imori of Hiroshima University for valuable comments on statistical analysis.

## Online Methods

### Collection of images of wheals and their morphological classification

Images of skin eruptions (wheals) were collected from 105 patients with CSU who visited the Department of Dermatology at Hiroshima University Hospital between January 1, 2000 and April 15, 2022. The institutional ethics review board approved the study protocol (Ethical Committee for Epidemiology of Hiroshima University, Hiroshima, Japan, approval number; E2020-2388), which involved analyzing images stored in the secured hospital database. Six dermatologists, specializing in allergic skin diseases, classified the morphology of wheals on each CSU patient into one of the following seven major categories: annular, broken-annular, geographic, circular, dot, uniform (shapeless and spread throughout the body), and none of the above (NA).

The uniform and NA cases were considered unclassifiable based on the EGe criteria; therefore, six categories were used in the final analysis. Sample number 105 that exceeded the minimal number required for an error with the true classification result within 8% and the significance level (*α*) was 5% (Thompson 1987).

### Statistical analysis

To classify the eruption patterns of patients, we used the vote-counting method, performed by six dermatologists. The reliability **SB** was calculated as follows:

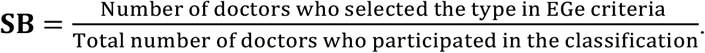

**SB** is the probability of *p*(*A* ∧ *B*)/*p*(*A*), where *p*(*A*) and *p*(*B*) are the probabilities that eruption patterns are classified in one of two classes or one of five types by doctors, respectively (Table 1). The reliability **SS** was calculated as follows:

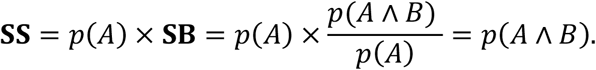

**SS** is the probability that both the class and type of eruption pattern were chosen correctly and that the **SS**-based analysis would provide more reliable statistical results. To confirm the statistical verification of the classifiable case data, we calculated the *P*-value as follows:

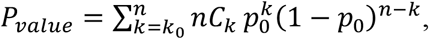

where, *k*_0_ is the sample number of classifiable cases, *n* is the total sample number, and *p*_0_ is the probability of the null/alternative hypothesis that the classifiable rate in the population is below/above *p*_0_%.

### Function representing the skin eruption state

To reflect the eruption state of the skin 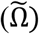 based on the molecular dynamics of the dermis (*Ω*_*D*_), we define the eruption state function, 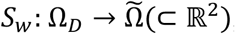), as follows:

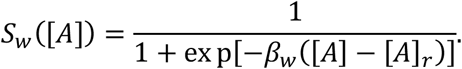

This function connected the dynamics of the coagulation factors ([*A*](**x**, t) ≡ [*C*](**x**, t)) leaked from blood vessel or histamine ([*A*](**x**, t) ≡ [*H*_*M*_](**x**, t)) released from mast cells in the dermis to the state of epidermal inflammation 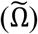. *β*_*w*_ is the positive value and [*A*]_*r*_ is the threshold value at which the eruption appears on the skin. This function allows for two states of eruption states: eruption present (1) or absent (0) (Seirin-Lee et al., 2020). Because the patterning dynamics of the histamine concentration of mast cells and coagulation factors were identical (Supplementary Fig. S1–S5), we plotted the representative results as [*A*](**x**, t) ≡ [*C*](**x**, t) in this study.

### Cell culture and impedance assay

Endothelial cell growth supplement (ECGS), bovine serum albumin (BSA), and lipopolysaccharide (LPS) were from Sigma-Aldrich Japan (Tokyo, Japan). The human umbilical vein endothelial cells (HUVECs) were obtained from American Type Culture Collection (ATCC, Manassas, VA, USA). Dulbecco’s modified Eagle’s medium (DMEM)/F12, penicillin, streptomycin, trypsin, and heparin were from Thermo Fisher Scientific (Waltham, MA, USA). Factor VIII-deficient human plasma and factor III (TF) were from COSMO BIO Co., Ltd. (Tokyo, Japan). Histamine was obtained from Wako Pure Chemical Industries, Ltd. (Osaka, Japan). HUVECs were cultured in DMEM/F12 supplemented with 10% fetal calf serum (FCS), 100 U/mL penicillin, 100 μg/mL streptomycin, 40 μg/mL ECGS, and heparin.

For the impedance analysis, HUVECs were harvested using trypsin and seeded onto E-Plates (Roche Applied Science, Upper Bavaria, Germany) at a density of 50,000 cells/well. On day 2, cells were treated with LPS (100 ng/mL) and histamine (10 μM) only for the TF stimulation assay. On day 3, the E-plate was set on an iCElligence microelectronic biosensor system (Roche Applied Science) and impedance termed “cell index (CI)” was measured every 10 s. For the TF stimulation assay (Fig. 2H), cells were stimulated with indicated TF concentrations in DMEM/F12 with 2% BSA and 1% factor VIII-deficient human plasma without any supplements and FCS. For the histamine stimulation assay (Fig. 2E), cells were stimulated with indicated concentrations of histamine in DMEM/F12 supplemented with 10% FCS.

## Supplementary Information Captions

### 1. Supplementary Notes

**1.1** Model description and method for numerical simulations

**1.2** Data conversion and inhibition functions

**1.3** Parameter estimation

**1.4** Modeling descriptions of histamine release in basophils and mast cells

**1.5** Leakage function estimation

**1.6** Statistical analysis for patients’ data

**1.7** Sensitivity analysis

**1.8** Simulation space and numerical method

### 2. Supplementary figures and tables

**Fig. S1:** Gap formation effect by tissue factor (TF) and histamine.

**Fig. S2-1, S2-2, S2-3, S2-4, S2-5**: Dynamics of histamine released from basophils ([*H*_*B*_]), tissue factor expressed on vascular endothelial cells ([*TF*]), coagulation factors leaked from blood vessel ([*C*]), and histamine released from mast cells ([*H*_*M*_]).

**Fig. S3:** Result for total index

**Table S1:** Details of parameter values for each wheal pattern used in Fig. 4 and representative parameter set for sensitivity analysis.

