## Supplementary Information for "Mathematical-structure based Morphological Classification of Skin Eruptions and Linking to the Pathophysiological State of Chronic Spontaneous Urticaria"

*Corresponding authors：

#### 1. Supplementary Notes

##### 1.1 Model description and method for numerical simulations

We recall the CSU model in the main text.

$$\frac{d[H_B]}{dt} = \delta_B + f^B_{activation}([TF])\chi_B(\mathbf{x},t)\left(1 - g^B_{inhibition}([H_B])\right) - \mu_B[H_B], on\ \Omega_B \quad (1a)$$

$$\frac{d[TF]}{dt} = \delta_T + f^T_{activation}([H_B],[H_M])\left(1 - g^T_{inhibition}([H_B],[H_M])\right) - \mu_T[TF], on\ \Omega_E \quad (1b)$$

$$\frac{\partial[C]}{\partial t} = D_c\nabla^2 C + f_{leakage}(\mathbf{x},t) - \mu_C[C], on\ \Omega_D \quad (1c)$$

$$\frac{\partial[H_M]}{\partial t} = D_M \nabla^2[H_M] + \delta_M + f_{activation}^M([C])\chi_M(\mathbf{x},t)\left(1 - g_{inhibition}^M([H_M])\right) - \mu_M[H_M], on\ \Omega_D \quad (1d)$$

The detailed equations which we used for numerical simulations are given to

$$\frac{d[H_B]}{dt} = \delta_B + \gamma_B[TF]\chi_B(\mathbf{x},t)\left(1 - \frac{\alpha_B[H_B]^2}{\alpha_{B0} + [H_B]^2}\right) - \mu_B[H_B], \quad on\ \Omega_B \quad (s1a)$$

$$\frac{d[TF]}{dt} = \delta_T + \frac{\gamma_T([H_B] + [H_M])}{\gamma_{T0} + [H_B] + [H_M]}\left(1 - \frac{\alpha_T([H_B] + [H_M])^2}{\alpha_{T0} + ([H_B] + [H_M])^2}\right) - \mu_T[TF], \ on\ \Omega_E \quad (s1b)$$

$$\frac{\partial[C]}{\partial t} = D_c \nabla^2 C + \frac{\gamma_C}{1 + \exp\left(-\beta([TF] - T_{sw})\right)} - \mu_C[C], \ on\ \Omega_D \quad (s1c)$$

$$\frac{\partial[H_M]}{\partial t} = D_M \nabla^2[H_M] + \delta_M + \gamma_M[C]\chi_M(\mathbf{x},t)\left(1 - \frac{\alpha_M[H_M]^2}{\alpha_{M0} + [H_M]^2}\right) - \mu_M[H_M], \ on\ \Omega_D \quad (s1d)$$

where $\chi_B(\mathbf{x},t)$ and $\chi_M(\mathbf{x},t)$ are defined by

$$\chi_B(\mathbf{x},t) = \begin{cases} 1 & if\ \int_0^t [H_B](\mathbf{x},\ t)dt \le H_B^{total} \\ 0 & otherwise \end{cases} \quad \text{on } \Omega_B,$$

$$\chi_M(\mathbf{x},t) = \begin{cases} 1 & if\ \int_0^t [H_M](\mathbf{x},\ t)dt \le H_M^{total} \\ 0 & otherwise \end{cases} \quad \text{on } \Omega_M.$$

$H_B^{total}$ and $H_M^{total}$ are the total amounts of histamine contained in basophils and mast cells, respectively. We assume that the total amount of histamine is limited because the time scale of eruption dynamics is from several minutes to hours, which is sufficiently shorter than the time scale of intracellular histamine reproduction (Hattori & Seifert R, 2017; Seirin-Lee, Yanase, et al., 2020).

We assume that the concentrations of histamine and TF are in stable equilibrium states before external stimulation. External stimuli are represented as spatially random perturbations near equilibrium. The mathematical formulation describing the initial conditions is given by

$$[H_B](\mathbf{x},0) = \frac{\gamma_B}{\mu_B}\left(1 + \epsilon\varphi_1(\mathbf{x})\right), \qquad [TF](\mathbf{x},0) = \frac{\gamma_T}{\mu_T}\left(1 + \epsilon\varphi_2(\mathbf{x})\right),$$

$$[H_M](\mathbf{x},0) = \frac{\gamma_M}{\mu_M}\left(1 + \epsilon\varphi_3(\mathbf{x})\right), \qquad [C](\mathbf{x},0) = 0,$$

where $\varphi_1(\mathbf{x}), \varphi_2(\mathbf{x})$, and $\varphi_3(\mathbf{x})$ are random valuable functions and $\epsilon$ is the scale of perturbation which we typically chose 0.01 (1% of equilibrium concentration).

Since we consider our model in the human body, we assumed zero-flux boundary conditions. To solve the main equations, we used C programming code written by the Alternating-Direction Implicit (ADI) method (Morton K.W. & Mayers D.F., 1994). The accuracy of the code was confirmed by regenerating examples from published papers and test simulations with smaller grids of time and space.

**1.2 Data conversion and inhibition functions**

To specify the detailed form of the inhibition rate functions to be included in the model $(s1a)$-$(s1d)$, we converted the adenosine concentration data of Fig.2A, B to histamine concentration data. Because the adenosine is released simultaneously with the release of histamine from basophils and hsMCs (Rudich et al., 2012), we assumed that the amount of released adenosine from mast cells or basophils is proportional to the amount of released histamine, namely,

$$[\text{Adenosine concentration }] = \kappa[\text{Histamin concentration}], \quad (s2)$$

where $\kappa$ is proportional coefficient. Using this equation, we converted the data of Fig.2A, B with histamine concentrations and estimated the specific forms of $g^{B}_{inhibition}$ and $g^{M}_{inhibition}$, as shown in Fig. 2C, by Hill functions such that

$$g^{B}_{inhibition}([H_B]) = \frac{\alpha_B [H_B]^2}{\alpha_{B0} + [H_B]^2},$$

$$g^{M}_{inhibition}([H_M]) = \frac{\alpha_M [H_M]^2}{\alpha_{M0} + [H_M]^2}$$

with parameter values, $\alpha_B$, $\alpha_{B0}$, $\alpha_M$, and $\alpha_{M0}$ (See the next section 1.3 for the estimation method in more detail). Similarly, we estimated the inhibition rate function of TF by using data conversion from Fig.2D to the Hill function (Fig.2E),

$$g^{T}_{inhibition}([H_B], [H_M]) = \frac{\alpha_T ([H_B] + [H_M])^2}{\alpha_{T0} + ([H_B] + [H_M])^2}.$$

**1.3 Parameter estimation**

**Estimation of parameters for the inhibition functions: $g^{B}_{inhibition}([H_B])$, $g^{M}_{inhibition}([H_M])$, $g^{T}_{inhibition}([H_B], [H_M])$**

The three inhibitory functions are similar in form. Thus, we here give an estimation method in general form. From the experimental data Figs.2A, 2B, and 2D, we first directly estimated the values of $\hat{\alpha}$ and $\tilde{\alpha}$ by using the following inhibition function of adenosine valuable $[A]$ such that

$$1-\frac{\hat{\alpha}[A]^2}{\tilde{\alpha}+[A]^2}.$$

Then, by instituting the equation $(s2)$ ($[A]=\kappa[H]$) into the above equation, we obtained

$$\frac{\hat{\alpha}[A]^2}{\tilde{\alpha}+[A]^2}=\frac{\hat{\alpha}[H]^2}{\frac{\tilde{\alpha}}{\kappa^2}+[H]^2}.$$

In here, we introduce non-dimensional scale of histamine concentration by $[\bar{H}]=[H]/H^*$. That is,

$$\frac{\hat{\alpha}[H]^2}{\frac{\tilde{\alpha}}{\kappa^2}+[H]^2}=\frac{\hat{\alpha}[\bar{H}]^2}{\frac{\tilde{\alpha}}{(\kappa H^*)^2}+[\bar{H}]^2}. \qquad (s3)$$

Next, we tested numerical simulations arbitrarily changing the value of $\kappa H^*$ and found the proper value at which the pattern reproduced as seen in patients with urticaria. We defined $\kappa H^*$ as a tuning parameter that links *in vitro* experimental data and *in silico* data. In simulations of this paper, $\kappa H^*$ was approximated to 89.0 with respect to the inhibition functions of $[H_B]$ and $[H_M]$, and $[TF]$. $\hat{\alpha}$ and $\tilde{\alpha}$ were estimated directly from *in vitro* experiments and given to $(\alpha_{B0}, \alpha_B)=(50/(\kappa H^*)^2, 0.335)$, $(\alpha_{M0}, \alpha_M)=(0.35/(\kappa H^*)^2, 0.865)$, $(\alpha_{T0}, \alpha_T)=(71400/(\kappa H^*)^2, 1.0)$ in the mathematical model (s1$a$)-($s1d$).

**Estimation of parameters for activation function: $f^{T}_{activation}([H_B],[H_M])$**

From *in vitro* experimental data of Fig. 2F, we estimated the activation function of TF to the function of histamine concentration $[H]$.

First, we rescaled the reaction of LPS and Histamine by

$$\sqrt{[H](nM)\times[LPS]\left(\frac{ng}{ml}\right)}\approx\sqrt{[H]^2\bar{\kappa}^2}=[H]\bar{\kappa}, \qquad (s4)$$

where we assumed $[LPS]\approx\bar{\kappa}^2[H]$ with a scaling parameter $\bar{\kappa}^2$ of the dimension $[\frac{ng}{ml\times nM}]$. Next, from the data of Fig. 2F, we estimated the function of TF expression with respect to the scaled value of $(s4)$ by

$$[\mathrm{TF-mRNA}]=\frac{b[H]\bar{\kappa}}{a+[H]\bar{\kappa}}=\frac{b[H]}{\frac{a}{\bar{\kappa}}+[H]} \qquad (s5)$$

using the least square method. The inferred parameter values are $a/\bar{\kappa}=220$ and $b=1690$.

Finally, introducing non-dimensional scales of histamine and TF concentrations by $[\bar{H}] = [H]/H^*$ and $[\overline{TF}] = [TF]/TF^*$, respectively, with respect to the equation $(s5)$, we had

$$f^T_{activation}([H_B],[H_M]) = \frac{\frac{b}{TF^*}[\bar{H}]}{\frac{a}{\kappa}H^* + [\bar{H}]}.$$

We chose the values of $H^*$ and $TF^*$ by testing simulations. We arbitrarily changed these values at which the pattern reproduced as seen in patients with urticaria.. In the representative simulations, we chose $H^* = 10^{-2}$ and $TF^*$= 402.38. Finally, we obtained $\gamma_T = 7.0, \gamma_{T_0} = 2.2$.

##### 1.4 Modeling descriptions of histamine release in basophils and mast cells: $f^B_{activation} = \gamma_B[TF]$ and $f^M_{activation} = \gamma_M[C]$

The degranulation of hsMCs and basophils is induced by the complement component C5a converted from C5, which is cleaved by coagulation factors triggered by TFs in a concentration-dependent manner (Yanase et al., 2021). Thus, the increasing rate of histamine released from basophil or histamine are written by

$$f^B_{activation} \ \ or \ \ f^M_{activation} = \tilde{\gamma}[\text{coagulation factors in intravascular/extravascular }],$$

where $\tilde{\gamma}$ is an increase ratio per unit time. Moreover, the amount of coagulation factors activated by TF in intravascular will be proportional to the expressed TF, namely: $[\text{coagulation factors in intravascular}] = \gamma[TF]$, thus we have $f^B_{activation} = \gamma\tilde{\gamma}[TF]$. Replacing parameter notations, we obtain that

$$f^B_{activation} = \gamma_B[TF] \text{ and } f^M_{activation} = \gamma_M[C],$$

where $\gamma_B$ is the histamine release rate of basophils by tissue factors and $\gamma_M$ is the histamine release rate of mast cells induced by the leaked coagulation factors.

##### 1.5 Leakage function estimation: $f_{leakage}(\mathbf{x},t)$

We approximated the increasing rate of leaked coagulation factors reflected in the mathematical model (1c) induced by intercellular gap formation driven by a TF-dependent switch-like role, as follows:

$$f_{leakage}(\mathbf{x},t) \propto \frac{d[\text{Area of gap formation}]}{dt}(\mathbf{x},t) \times \chi_{sw}([TF](\mathbf{x},t))$$

where $\chi_{sw}$ is a TF-dependent switch-like function.

First, we inferred the TF-dependent switch-like function ($\chi_{sw}$) by calculating the relative CI, $(1 - CI)/(1 - CI_{100\,ng/ml})$, which gives the relative variation in the size of the CI for each time point. We calculated the average relative CI at [0.2, 0.6] (hour) for each TF concentration and found the switch-like function as shown in Fig. 2I. Based on the result, we propose that the gap formation variation could be described by a step function, and we defined $\chi_{sw}$ as follows (Fig.2J).

$$\chi_{sw}([TF]) = \frac{1}{1 + \exp\left[-\beta([TF] - T_{sw})\right]}$$

where $T_{sw}$ is a threshold value of TF at which a cell gap formation is formed, and $\beta$ is a positive constant expressing the stiffness of switching.

Next, to infer the function of $d[\text{Area of gap formation}]/\text{dt}$, we used the CI data of various TF and histamine concentrations (Fig.S1A and C) after the gap was formed because the gap area is proportional to the level of CI, and we obtain the following equation as shown in Fig.S1B and D:

$$\frac{d}{dt}[\text{Area of gap formation}](\cdot, t) \propto \frac{\Delta[CI](t)}{\Delta t} \approx 0.$$

Thus, we obtain $[\text{Area of gap formation}](\cdot, t) \approx \gamma_{TF} + \gamma_{Histamine} = \gamma_C$ (constant). This estimation indicates that the gap formation may quickly approach the size limit, independent of TF and histamine concentrations if the TF state is switched on. Thus, the TF switch-like dynamics may play a key role in onset/no-onset of CSU.

##### 1.6 Statical analysis for patients' data

**Verification of total sample number for classification**

We classified the eruption types into six categories: annual type, broken-annual type, geographic type, circular type, dot type, unclassifiable type. Based on the statical analysis of (Thompson, 1987), the minimal number of samples required for the error with the true classification result to be within 8 % with the significance level ($\alpha$) of 5 % is given to 94 samples. The number of total samples which we used is 105 patient samples.

**Verification of classifiable ratio 87.6 %**

To confirm that the classifiable ratio 87.6% is not rare event with respect to numerous sample data, we used P-value test. Let us assume that the null hypothesis ($H_0$); The classifiability of the population by EGe Criteria is no higher than 79%, and the alternative hypothesis ($H_1$); The classifiability of the population by EGe Criteria exceeds 79%. We then obtain $P_{value} = \sum_{k=k_0}^{n} nC_k \, p_0^k (1 - p_0)^{n-k} < 0.016$ where $k_0 = 92, n = 105,$ and $p_0 = 0.79$. Thus, we adopt $H_1$ depending on the significance level ($\alpha$=2.5 %) of the one-tailed test.

##### 1.7 Sensitivity analysis

For the sensitivity analysis, we used the variance-based sensitivity analysis by using Extended Fourier Amplitude Sensitivity Test (eFAST) Method (See Appendix A.3 of (Seirin-Lee, Gaffney, et al., 2020) for the details of the method). For the calculations of the first order sensitivity index and the total-effect index, we used the frequencies {19, 23, 59, 77, 91, 107, 113, 121, 125, 133, 149, 157, 161} and {1, 1, 3, 5, 7, 9, 11, 13, 15, 17, 19, 21, 168}, respectively. Thus, the total sample size, $N_s = (2M\omega_{max} + 1)N_r$, is given to 1289 and 1345, respectively, with $M = 4, N_r = 2$.

We calculated two sensitivity measures, the first order index and total-effect index. The first order index indicates the influence of parameter $p_i$ on the variance of each feature measure (the model output), independent of interactions with the other parameters. The total-effect index indicates the effect of parameter/network $p_i$ when interactions with the other parameters/networks are included. These two measures give a full quantification of the importance of parameter/network $p_i$ and whether the extent whether this is a direct influence or through interactions with other parameters/networks or both. We show the representative result of the first order index in Fig.6A-E. The similar results are shown in the total-effect index (Fig. S2).

Based on each feature of wheal patterns in Table 2, we defined the sensitivity function ($S_f$) for each type of eruption patterns as the following.

**[Boundary: KF1]**

We defined the sensitivity function which give how much the boundary property is maintained, as the following.

When

$$Bd \equiv \left| \int_0^L \int_0^L |S_w(x,y,t^*) - S_w(x,y,t_0)| dxdy - \int_0^L \int_0^L |S_w(x,y,t^*)| dxdy - \int_0^L \int_0^L |S_w(x,y,t_0)| dxdy \right| < \mathcal{E},$$

$$S_f = \left| \int_0^L S_w\left(x, \frac{L}{2}, t^*\right) - S_w\left(x, \frac{L}{2}, t_0\right) dx \right|.$$

**[Boundary-disconnected: KF2]**

We defined the sensitivity function which give how much the disconnected property of boundary wheal is promoted, as the following.

When $Bd < \varepsilon$,

$$S_f = \frac{1}{2}\left\{\left|\int_0^L \int_0^{L/2} S_w(x,y,t^*)dxdy - \int_0^L \int_{L/2}^L S_w(x,y,t^*)dxdy\right| + \left|\int_0^L \int_0^{L/2} S_w(x,y,t^*)dydx - \int_0^L \int_{L/2}^L S_w(x,y,t^*)dydx\right|\right\}$$

**[Area: KF3]**

We defined the sensitivity function which give how much the area property is maintained, as the following.

When

$$Ar \equiv \int_0^L \int_0^L |S_w(x,y,t^*) - S_w(x,y,t_0)| - \int_0^L \int_0^L |S_w(x,y,t^*)| < 0,$$

$$S_f = \left|\int_{\frac{L}{2}-\delta_\varepsilon}^{\frac{L}{2}+\delta_\varepsilon} \int_{\frac{L}{2}-\delta_\varepsilon}^{\frac{L}{2}+\delta_\varepsilon} S_w(x,y,t^*)dxdy - \int_{\frac{L}{2}-\delta_\varepsilon}^{\frac{L}{2}+\delta_\varepsilon} \int_{\frac{L}{2}-\delta_\varepsilon}^{\frac{L}{2}+\delta_\varepsilon} S_w(x,y,t_0)dxdy\right| \frac{1}{t^* - t_0}$$

where $\delta_\varepsilon$ is a small radius.

**[Area fusion: KF4]**

To determine the extent to which area-type weals fuse, we defined a sensitivity function that gives the degree of area weal expansion as follows.

When $Ar < 0$,

$$S_f = \frac{1}{t^* - t_0} \sum_{t=t_0}^{t^*} \frac{1}{\Delta t} \left|\int_0^L \int_0^L S_w(x,y,t+\Delta t)\, dxdy - \int_0^L \int_0^L S_w(x,y,t)dxdy\right|$$

**[Punctate: KF5]**

We defined the sensitivity function which give how much the small and fixed size wheal is maintained, as the following.

When

$$Dot \equiv \int_0^L \int_0^L S_w(x,y,t^*)\, dxdy < \varepsilon_0,$$

$$S_f = \sum_{t=0}^{t^*} \int_0^L \int_0^L S_w(x,y,t)\,dxdy.$$

To calculate the first order sensitivity index and the total-effect index, we chose initial conditions by which a wheal pattern emerges in the center area of simulation space. The example is shown in Fig. S2G. The results of total index are shown in Fig. S2A-F. In the case of the total-effect index of broken-boundary, the index value of dummy parameter was higher than all the other model parameters, so that we could not obtain any valid information. Nonetheless, we found that the effect of the positive networks for histamine release from basophils ($\gamma_B, \delta_B$) are negligible, indicating that basophils may not play an important role in the variation of wheal shapes and be involved in the triggering phase of CSU development.

##### 1.8 Simulation space and numerical method

We simulated a two-dimensional (2D) model of the dermal region ($\Omega_D$) and vascular endothelial tissue ($\Omega_E$), whereas $\Omega_B(\subset \Omega_E)$ is a random region of vascular endothelial tissue where histamine released from basophils affected endothelial cells. In model simulations, we set $\Omega_D = \Omega_E = [0,L] \times [0,L]$ and $\Omega_B = \{\mathbf{x} | \mathbf{x} = \mathrm{RAND}([0,L] \times [0,L])\}$, where RAND is the random value function of the uniform distribution. The numerical simulation code was written using C language based on the alternating-direction implicit (ADI) method.

### 2 Supplementary figures and tables

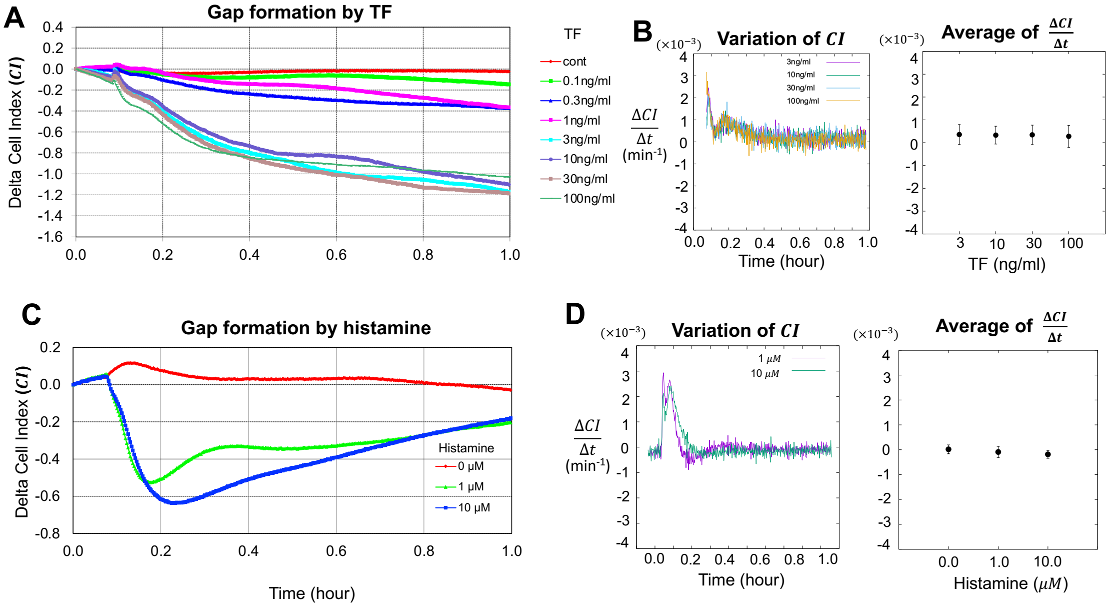


**Fig.S1: Gap formation effect by Tissue factor (TF) and histamine.** (A) Temporal variation in cell index (CI) at different TF concentrations. (B) Quantitative analysis of variation in CI at different TF concentrations. (C) Temporal variation in CI with varying histamine concentrations. Endothelial cells were stimulated with histamine at indicated concentrations in DMEM/F12 supplemented with 10% fetal calf serum (FCS). (D) Quantitative analysis of CI variation with histamine concentrations.

(Fig. S2-1) Annular pattern

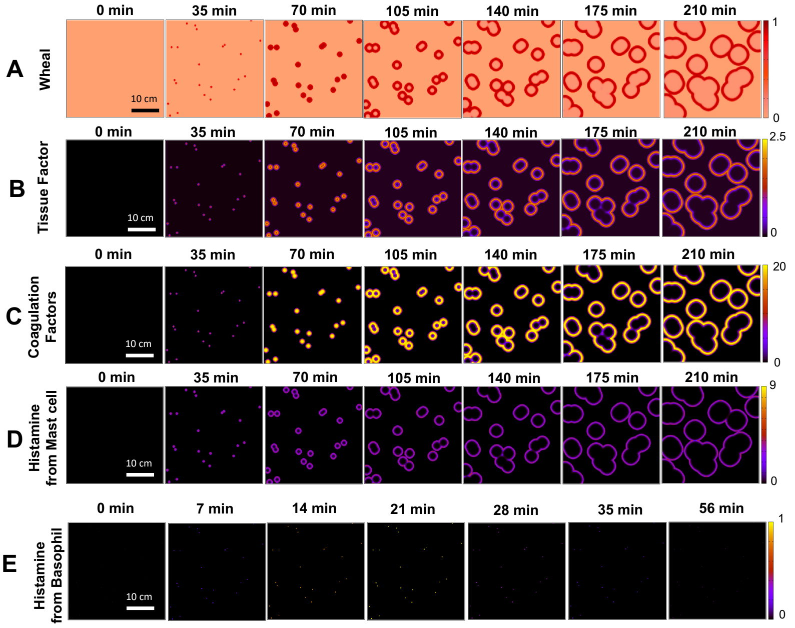


(Fig.S2-2) Broken Annular pattern

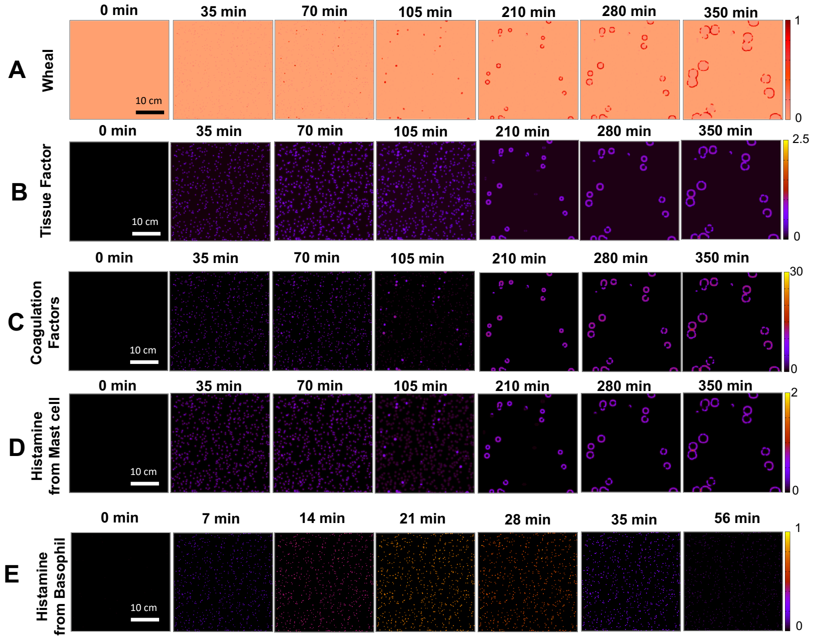

(Fig.S2-3) Geographic pattern

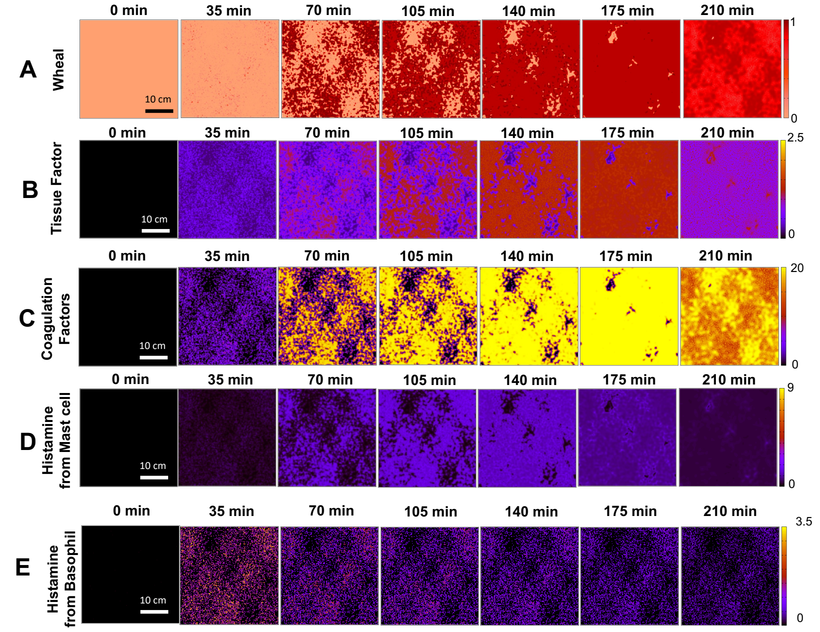


(Fig.S2-4) Circular pattern

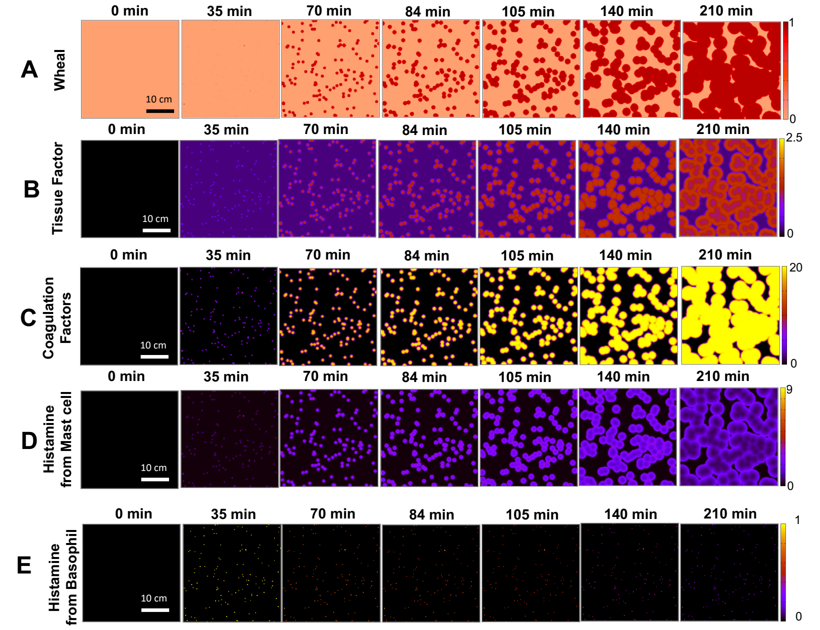

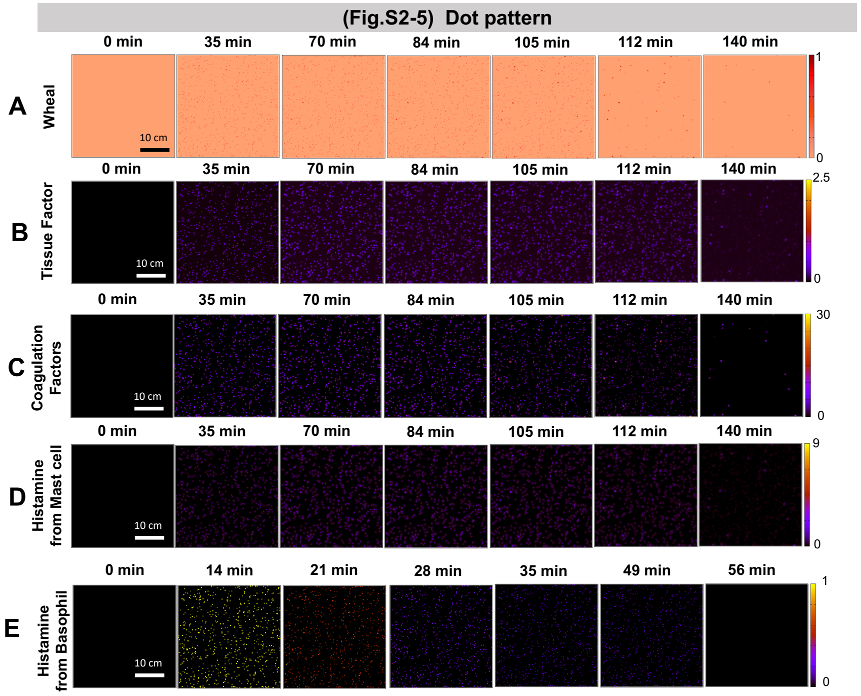


**Fig.S2-1, S2-2, S2-3, S2-4, S2-5**: Dynamics of wheal patterns on skin (A), tissue factor expressed on vascular endothelial cells ($[TF]$) (B), coagulation factors leaked from blood vessel ($[C]$) (C), histamine released from mast cells ($[H_M]$) (D), and histamine released from basophils ($[H_B]$) (E) for each type of pattern.

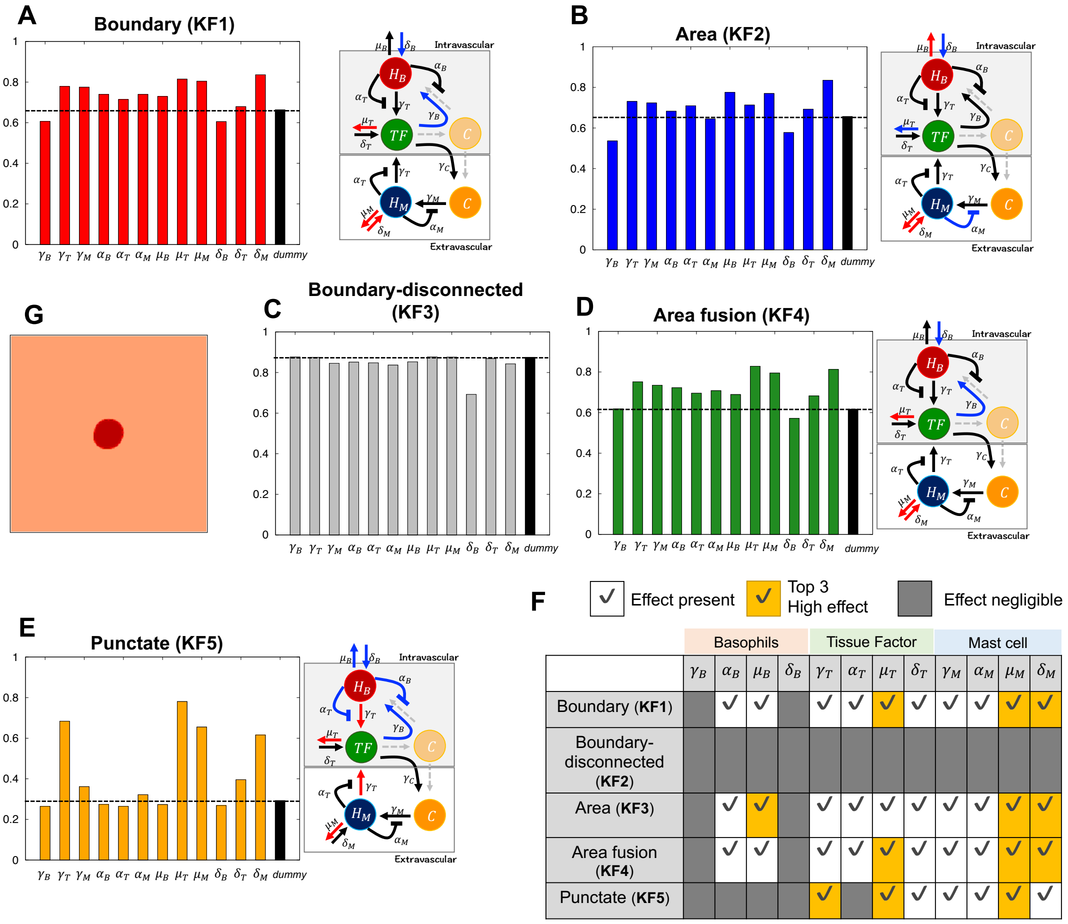


| | Basophils | | | | Tissue Factor | | | | Mast cell | | | |
|---|---|---|---|---|---|---|---|---|---|---|---|---|
| | $\gamma_B$ | $\alpha_B$ | $\mu_B$ | $\delta_B$ | $\gamma_T$ | $\alpha_T$ | $\mu_T$ | $\delta_T$ | $\gamma_M$ | $\alpha_M$ | $\mu_M$ | $\delta_M$ |
| Boundary (**KF1**) | | ✓ | ✓ | | ✓ | ✓ | ✓ | ✓ | ✓ | ✓ | ✓ | ✓ |
| Boundary-disconnected (**KF2**) | | | | | | | | | | | | |
| Area (**KF3**) | | ✓ | ✓ | | ✓ | ✓ | ✓ | ✓ | ✓ | ✓ | ✓ | ✓ |
| Area fusion (**KF4**) | | ✓ | ✓ | | ✓ | ✓ | ✓ | ✓ | ✓ | ✓ | ✓ | ✓ |
| Punctate (**KF5**) | | | | | ✓ | | ✓ | ✓ | ✓ | ✓ | ✓ | ✓ |

**Fig.S3: Result for the total-effect index (A-F) and the example simulation used for the sensitivity analysis (G).** The representative parameter value set in Table S1 was used for analysis. The red/blue arrows indicate a high/negligible effect. Since none of the total-effect indices were markedly different from the dummy parameter case for KF1-KF5, the arrows for the top three total-effect indices are indicated in red.

**Table S1. The representative parameter value set for sensitivity analysis and the details of parameter values for each eruption pattern used in simulations.** Parameter values estimated from the experimental data in Fig. 2 is noted by *. Dimension values are shown in parentheses. [ ] represents the parameter range which used for sensitivity analysis.

| Parameters | Representative Value Set | Annular | Broken annular |
|---|---|---|---|
| Spatial length ($L \times L$) | $1 \times 1$ ($35.5 \times 35.5\text{cm}^2$) | $1 \times 1$ | $1 \times 1$ |
| Time scale ($t$) | 1.0 (420 sec) | 1.0 | 1.0 |
| Diffusion rates of histamine ($D_M$) and coagulation factors ($D_C$) | $4.7 \times 10^{-6}$ ($1.412 \times 10^{-5}$ $\text{cm}^2/\text{sec}$)* | $4.7 \times 10^{-6}$ ($1.412 \times 10^{-5}$ $\text{cm}^2/\text{sec}$) | $4.7 \times 10^{-6}$ ($1.412 \times 10^{-5}$ $\text{cm}^2/\text{sec}$) |
| Basal secretion rates of histamine from basophil ($\delta_B$) | [0, 1.0] ([0.0, 0.0024] sec$^{-1}$) | 0.1 (0.00024 sec$^{-1}$) | 0.1 (0.00024 sec$^{-1}$) |
| Histamine release rate of basophil ($\gamma_B$) | [0, 5.0] ([0.0, 0.012] sec$^{-1}$) | 5.0 (0.012 sec$^{-1}$) | 5.0 (0.012 sec$^{-1}$) |
| Parameter affecting the gradient of inhibition rate of histamine from basophil ($\alpha_{B0}$) | 0.00625 * | 0.00625 * | 0.00625 * |
| Maximal inhibition rate of histamine from basophil ($\alpha_B$) | [0, 1.0] | 0.335 * | 0.335 * |
| Basal decay rate of histamine of basophil ($\mu_B$) | [0, 1.0] ([0.0, 0.0024] sec$^{-1}$) | 1.0 (0.0024 sec$^{-1}$) | 1.0 (0.0024 sec$^{-1}$) |
| Basal secretion rates of tissue factor ($\delta_T$) | [0, 1.0] ([0.0, 0.0024] sec$^{-1}$) | 0.01 (0.000024 sec$^{-1}$) | 0.01 (0.000024 sec$^{-1}$) |
| Maximal increase rate of tissue factor ($\gamma_T$) | 4.2 * [0, 5.0] ([0.0, 0.012] sec$^{-1}$) | 7.0 (0.017 sec$^{-1}$) | 4.2 * (0.01 sec$^{-1}$) |
| Parameter determining the curve of increasing tissue factor ($\gamma_{T0}$) | 2.2 * | 2.2 * | 2.2 * |
| Basal decay rate of tissue factor ($\mu_T$) | [0, 1.0] ([0.0, 0.0024] sec$^{-1}$) | 1.0 (0.0024 sec$^{-1}$) | 1.0 (0.0024 sec$^{-1}$) |
| Parameter affecting the gradient of inhibition rate of tissue factor ($\alpha_{T0}$) | 8.925 * | 8.925 * | 8.925 * |
| Maximal inhibition rate of histamine from | 1.0 * [0, 1.0] | 1.0 * | 1.0 * |

| | | | |
|---|---|---|---|
| basophil ($\alpha_T$) | | | |
| Leakage rate of coagulation factors from blood vessel ($\gamma_C$) | 20.0 (0.0476 sec$^{-1}$) | 20.0 (0.0476 sec$^{-1}$) | 20.0 (0.0476 sec$^{-1}$) |
| Parameter determining the stiffness of switching ($\beta$) | 20.0 | 20.0 | 20.0 |
| Switch value of tissue factor ($T_{sw}$) | 0.67 | 0.67 | 0.67 |
| Basal decay rate of coagulation factors ($\mu_C$) | [0, 1.0] ([0.0, 0.0024] sec$^{-1}$) | 1.0 (0.0024 sec$^{-1}$) | 1.0 (0.0024 sec$^{-1}$) |
| Basal secretion rates of histamine from mast cell ($\delta_M$) | [0, 1.0] ([0.0, 0.0024] sec$^{-1}$) | 0.01 (0.000024 sec$^{-1}$) | 0.01 (0.000024 sec$^{-1}$) |
| Histamine release rate of mast cell ($\gamma_M$) | [0, 5.0] ([0.0, 0.012] sec$^{-1}$) | 5.0 (0.012 sec$^{-1}$) | 0.8 (0.002 sec$^{-1}$) |
| Parameter affecting the gradient of inhibition rate of histamine from mast cell ($\alpha_{M0}$) | $4.37\times 10^{-5}$ * | $4.37\times 10^{-5}$ * | $4.37\times 10^{-5}$ * |
| Maximal inhibition rate of histamine from basophil ($\alpha_M$) | 0.865 * [0, 1.0] | 0.865 * | 0.865 * |
| Basal decay rate of histamine from mast cell ($\mu_M$) | [0, 1.0] ([0.0, 0.0024] sec$^{-1}$) | 1.0 (0.0024 sec$^{-1}$) | 1.0 (0.0024 sec$^{-1}$) |
| Total amount of histamine of basophil ($H_B^{total}$) | 600 | 600 | 600 |
| Total amount of histamine of mast cell ($H_M^{total}$) | 600 | 600 | 600 |
| Parameter determining the stiffness of wheal state function ($\beta_w$) | 1.0 | 1.0 | 1.0 |

| **Parameters** | **Geographic** | **Circular** | **Dot** |
|---|---|---|---|
| Spatial length ($L \times L$) | $1 \times 1$ ($35.5 \times 35.5\text{cm}^2$) | $1 \times 1$ ($35.5 \times 35.5\text{cm}^2$) | $1 \times 1$ ($35.5 \times 35.5\text{cm}^2$) |
| Time scale ($t$) | 1.0 (420 sec) | 1.0 (420 sec) | 1.0 (420 sec) |
| Diffusion rates of histamine ($D_M$) and coagulation factors ($D_C$) | $4.7 \times 10^{-6}$ ($1.412 \times 10^{-5}$ $\text{cm}^2/\text{sec}$) | $4.7 \times 10^{-6}$ ($1.412 \times 10^{-5}$ $\text{cm}^2/\text{sec}$) | $4.7 \times 10^{-6}$ ($1.412 \times 10^{-5}$ $\text{cm}^2/\text{sec}$) |
| Basal secretion rates of histamine from basophil ($\delta_B$) | 0.1 (0.00024 sec$^{-1}$) | 0.1 (0.00024 sec$^{-1}$) | 0.5 (0.0012 sec$^{-1}$) |
| Histamine release rate of basophil ($\gamma_B$) | 5.0 (0.012 sec$^{-1}$) | 5.0 (0.012 sec$^{-1}$) | 20.0 (0.048 sec$^{-1}$) |

| | | | |
|---|---|---|---|
| Parameter affecting the gradient of inhibition rate of histamine from basophil ($\alpha_{B0}$) | 0.00625 * | 0.00625 * | 0.00625 * |
| Maximal inhibition rate of histamine from basophil ($\alpha_B$) | 0.335 * | 0.335 * | 0.335 * |
| Basal decay rate of histamine of basophil ($\mu_B$) | 1.0 (0.0024 $sec^{-1}$) | 1.0 (0.0024 $sec^{-1}$) | 1.0 (0.0024 $sec^{-1}$) |
| Basal secretion rates of tissue factor ($\delta_T$) | 0.01 (0.000024 $sec^{-1}$) | 0.01 (0.000024 $sec^{-1}$) | 0.01 (0.000024 $sec^{-1}$) |
| Maximal increase rate of tissue factor ($\gamma_T$) | 4.2 * (0.01 $sec^{-1}$) | 4.2 * (0.01 $sec^{-1}$) | 4.2 * (0.01 $sec^{-1}$) |
| Parameter determining the curve of increasing tissue factor ($\gamma_{T0}$) | 2.2 * | 2.2 * | 2.2 * |
| Basal decay rate of tissue factor ($\mu_T$) | 1.0 (0.0024 $sec^{-1}$) | 1.0 (0.0024 $sec^{-1}$) | 1.0 (0.0024 $sec^{-1}$) |
| Parameter affecting the gradient of inhibition rate of tissue factor ($\alpha_{T0}$) | 8.925 * | 8.925 * | 8.925 * |
| Maximal inhibition rate of histamine from basophil ($\alpha_T$) | 1.0 * | 1.0 * | 1.0 * |
| Leakage rate of coagulation factors from blood vessel ($\gamma_C$) | 20.0 (0.0476 $sec^{-1}$) | 20.0 (0.0476 $sec^{-1}$) | 20.0 (0.0476 $sec^{-1}$) |
| Parameter determining the stiffness of switching ($\beta$) | 20.0 | 20.0 | 20.0 |
| Switch value of tissue factor ($T_{sw}$) | 0.67 | 0.67 | 0.67 |
| Basal decay rate of coagulation factors ($\mu_C$) | 1.0 | 1.0 | 1.0 |
| Basal secretion rates of histamine from mast cell ($\delta_M$) | 0.01 (0.000024 $sec^{-1}$) | 0.01 (0.000024 $sec^{-1}$) | 0.01 (0.000024 $sec^{-1}$) |
| Histamine release rate of mast cell ($\gamma_M$) | 0.2 (0.00048 $sec^{-1}$) | 1.0 (0.0024 $sec^{-1}$) | 1.0 (0.0024 $sec^{-1}$) |
| Parameter affecting the gradient of inhibition rate of histamine from mast cell ($\alpha_{M0}$) | $4.37\times 10^{-5}$ * | $4.37\times 10^{-5}$ * | $4.37\times 10^{-5}$ * |
| Maximal inhibition rate of histamine from basophil ($\alpha_M$) | 0.865 * | 0.865 * | 0.865 * |
| Basal decay rate of | 0.1 | 0.1 | 0.1 |

| | | | |
|---|---|---|---|
| histamine from mast cell ($\mu_M$) | (0.00024 $sec^{-1}$) | (0.00024 $sec^{-1}$) | (0.00024 $sec^{-1}$) |
| Total amount of histamine of basophil ($H_B^{total}$) | 600 | 600 | 600 |
| Total amount of histamine of mast cell ($H_M^{total}$) | 600 | 600 | 600 |
| Parameter determining the stiffness of wheal state function ($\beta_w$) | 1.0 | 1.0 | 1.0 |
